# Polar Vantage and Oura physical activity and sleep trackers: A validation and comparison study

**DOI:** 10.1101/2020.04.07.20055756

**Authors:** André Henriksen, Frode Svartdal, Sameline Grimsgaard, Gunnar Hartvigsen, Laila Hopstock

## Abstract

**Background:** Consumer-based activity trackers are increasingly used in research as they have potential to increase activity participation and can be used for estimating physical activity. However, the accuracy of newer consumer-based devices is mostly unknown, and validation studies are needed.

**Objective:** The objective of this study was to test the accuracy of the Polar Vantage watch and Oura ring activity trackers for measuring physical activity, total energy expenditure, resting heart rate, and sleep duration, in free-living adults.

**Methods:** Twenty-one participants wore two consumer-based activity trackers (Polar, Oura), an ActiGraph accelerometer, an Actiheart accelerometer and heart rate monitor, and completed a sleep diary for up to seven days. We assessed Polar and Oura validity and comparability for physical activity, total energy expenditure, resting heart rate (Oura), and sleep duration. We analysed repeated measures correlation, Bland-Altman plots, and mean absolute percentage error.

**Results:** Polar and Oura were both strongly correlated (p<0.001) with ActiGraph for steps (Polar *r* 0.75, 95% CI 0.54-0.92. Oura *r* 0.77, 95% CI 0.62-0.87), moderate-to-vigorous physical activity (Polar *r* 0.76, 95% CI 0.62-0.88. Oura *r* 0.70, 95% CI 0.49-0.82), and total energy expenditure (Polar *r* 0.69, 95% CI 0.48-0.88. Oura *r* 0.70, 95% CI 0.51-0.83) and strongly or very strongly correlated (p<0.001) with the sleep diary for sleep duration (Polar *r* 0.74, 95% CI 0.56-0.88. Oura *r* 0.82, 95% CI 0.68-0.91). Oura had a very strong correlation (p<0.001) with Actiheart for resting heart rate (*r* 0.9, 95% CI 0.85-0.96). However, all confidence interval ranges were wide and mean absolute percentage error was high for all variables, except Oura sleep duration (10%) and resting heart rate (3%) where Oura under-reported on average one beat per minute.

**Conclusions:** Oura can potentially be used as an alternative to Actiheart to measure resting heart rate. For sleep duration, Polar and Oura can potentially be used as a replacement for a manual sleep diary, depending on acceptable error. Neither Polar nor Oura can replace ActiGraph for measuring steps, moderate-to-vigorous physical activity, and total energy expenditure, but may be used as an additional source of physical activity in some settings.

## Introduction

In a research setting, accelerometers are often used to objectively measure physical activity and related variables. Device output is converted into various estimates for physical activity, energy expenditure, sleep, and heart rate (for devices with heart rate sensor). A wide range of devices exist, aimed at both research [1] and the consumer marked [2].

Consumer-based activity trackers are increasingly used in research as they have potential to increase activity participation [3] and can be used for estimating physical activity and related variables [2, 4, 5]. Compared to research-based accelerometers and heart rate sensor, consumer-based activity trackers are often cheaper, less intrusive, and have increased battery and storage capacity. They can also decrease participant burden compared with self-report instruments such as physical activity and sleep diaries. Sleep diaries [6] are less resource demanding compared with polysomnography (PSG), but compliance can be challenging [7]. Using consumer-based activity trackers to measure sleep duration can therefore be a potential replacement for sleep diaries.

The accuracy of newer consumer-based devices is mostly unknown, and validation studies are needed. Validity of consumer-based activity trackers can be studied by comparison against research-based accelerometers (e.g. ActiGraph) that, in turn, have been validated against gold standard methods. The Polar Vantage watch and the Oura ring are two new consumer-based activity trackers that potentially can replace research-based accelerometers or self-report tools.

A previous lab-based validation study of Polar Vantage found that the accuracy of energy expenditure was moderately or higher, depending on performed activity type [8]. No validation or comparison study on physical activity or sleep has been conducted on the Polar Vantage to date, and no study on this activity tracker has been done in free-living populations. Similarly, de Zambotti et al. [9] conclude that the Oura shows “promising results” for sleep detection, when compared with PSG. However, there are no previous validation studies testing the validity of physical activity or energy expenditure for the Oura.

The aim of this study on free-living adults was therefore to test the validity and comparability of physical activity and energy expenditure, measured by Polar Vantage and Oura, compared to ActiGraph, as well as resting heart rate (RHR) measured by the Oura compared to Actiheart electrocardiograms (ECG). In addition, we compared sleep duration between a sleep diary and output from the Polar Vantage and Oura.

## Methods

### Instruments

The *Polar Vantage* activity tracker (Polar Electro oy, Finland) was released in 2018 and is equipped with a 50 Hz (i.e. measurements per second) triaxial accelerometer for physical activity tracking. It weighs 45-66 grams, has one week of battery life, and comes in multiple strap- and metal casing colours. Polar Vantage is a multisport watch to be worn on the wrist.

The *Oura* activity- and sleep ring (Oura oy, Finland) was released in 2018 and equips a 50 Hz triaxial accelerometer for physical activity tracking and a two infrared LED (light-emitting diode) photoplethysmograph for optical pulse measurements. It comes in sizes US 6 to US 13, weighs 4-6 grams, has six days of battery life, and comes in different shapes and colours. Oura is a smart ring to be worn on the finger and focuses on sleep and wellbeing by combining physical activity and heart rate parameters.

The *ActiGraph wGT3X-BT* (ActiGraph, Pensacola, FL, USA) is a triaxial accelerometer. Sample rate can be set to 30-100 Hz. It weighs 19 grams and has up to 25 days of battery life. It is extensively used to estimate activity in free-living research, as it provides reasonable estimates for physical activity intensity [10], steps [11], and energy expenditure [12].

The *Actiheart 4 (*CamNtech Ltd, Cambridge, UK) records heart rate using a 1-lead ECG, with 128 Hz sampling rate. It weighs less than 10 grams and is attached to the chest using two standard ECG electrodes. Actiheart is valid and reliable for heart rate detection [13].

The sleep diary contained a sub-set of questions from the Consensus Sleep Diary [14] relevant for sleep duration measured as time in bed; specifically question 1) “what time did you get into bed?”, and question 7) “what time did you get out of bed for the day?”.

### Participants and procedure

We used convenience sampling to recruit 21 participants. Participants were eligible for inclusion if they were above 18 years of age, had normal physical function, and were willing to wear all four devices and keep a sleep diary for five days. Participants were recruited among university students and staff, and people close to these participants.

We initialized devices with self-reported information on height, weight, age, sex, and dominant hand. Participants wore the Polar Vantage and Oura on the non-dominant hand. The ActiGraph was setup for 100 Hz recording and placed on the right hip, attached with an elastic band. The Actiheart was placed at the level of the fifth intercostal space at the sternum (medial part) and to the left (lateral part), attached with two 3M Red Dot 2238 electrodes (3M, St Paul, MN, USA). Participants were asked to wear all four devices simultaneously and complete the sleep diary for five days. Since devices were placed on participants on day one and removed on day five, expected valid days per person was up to three days. We used both an ActiGraph and an Actiheart, because neither device could record all variables (i.e. ActiGraph could not record heart rate and Actiheart could not record steps). Data were collected May-June 2019. Figure 1 show device placements.

**Figure 1.**
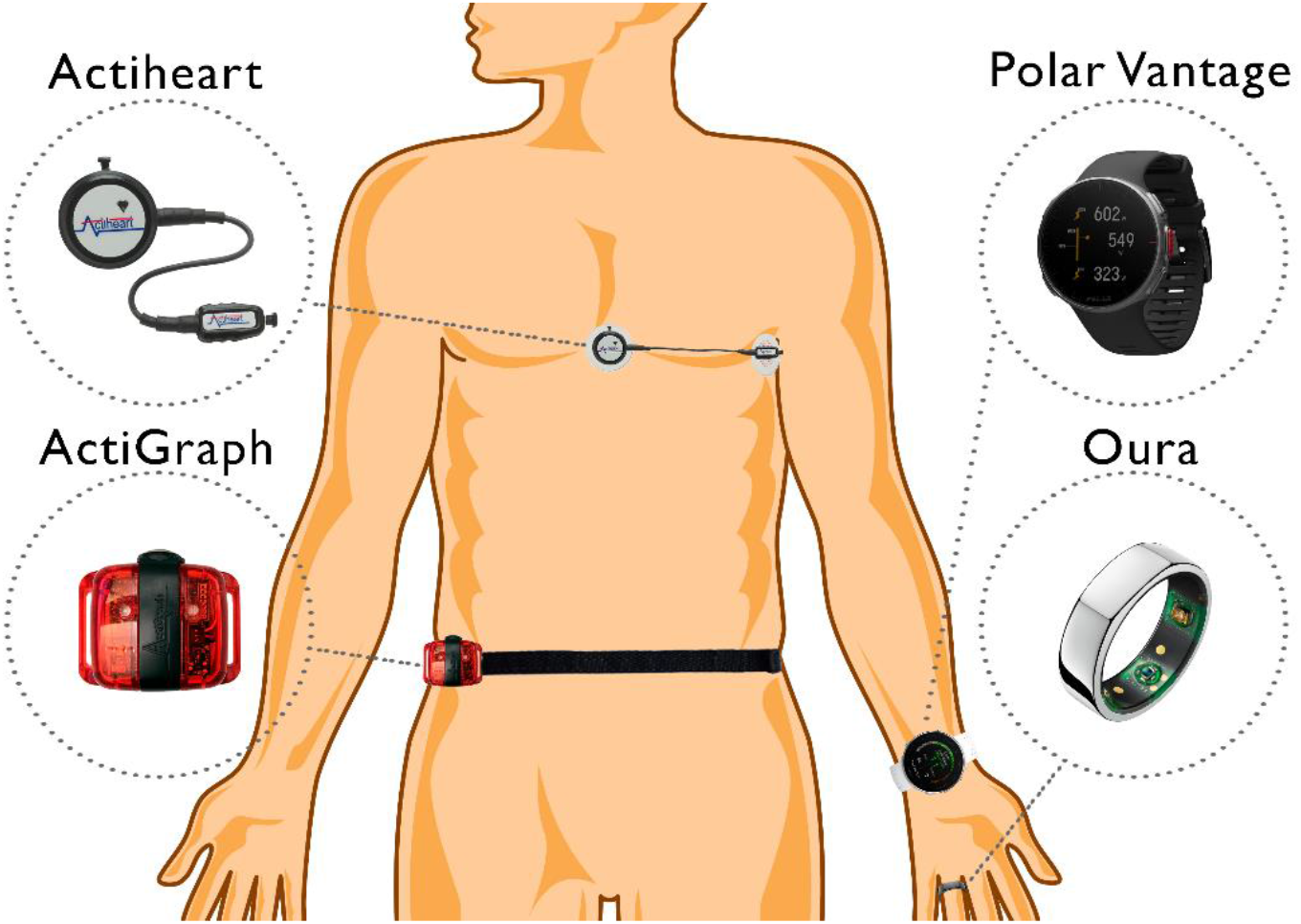
Illustration of instrument placement for Actiheart, Polar Vantage, ActiGraph, and Oura

An overview of instrument details and software used for setup, data download, and variable generation, is given in Table 1.

**Table 1.**
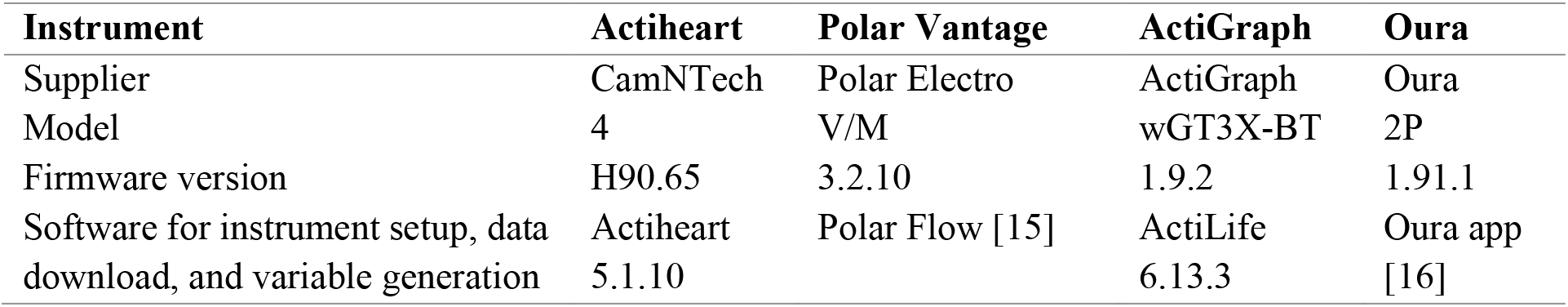
Overview of instrument supplier, model, firmware version, and software version used to setup, download, and generate output variables.

### Variable generation

We used Polar Flow (Polar Electro oy, Finland) to download daily Polar Vantage variables for steps, MVPA, TEE, and sleep duration (sleep time). The Oura application was used to download daily Oura variables for steps, MVPA, TEE, sleep duration (total sleep), and RHR. We used ActiLife (ActiGraph, Pensacola, FL, USA) to download ActiGraph accelerometer data and generate variables. We used triaxial activity (i.e. vector magnitude (VM)) counts to generate physical activity and energy expenditure variables, in addition to steps, which was reported directly. We calculated minutes of MVPA using cut-points defined by Sasaki et al. [10], i.e. >2690 VM. Activity energy expenditure was calculated using “Freedson VM3 combination 2011 + Williams work-energy equation” (Sasaki et al. [10]) and converted to TEE using the Schofield equation [17] for resting energy expenditure and removing 10% to account for diet-induced thermogenesis. The Actiheart software was used to download sleeping (i.e. resting) heart rate from the Actiheart (CamNtech Ltd, Cambridge, UK). Sleep duration (time in bed) was calculated based on manual recorded times (i.e. time-out-of-bed minus time-into-bed) from the sleep diary. We did not analyse sleep duration using the Actiheart software, as this feature is considered experimental.

We only included valid days in the analysis, defined as at least 10 hours of wear time [18]. Wear time for the ActiGraph was analysed using the ActiLife default settings for the Troiano [19] wear time algorithm. Wear time algorithms for Polar Vantage and Oura are unknown.

### Statistics

The ActiGraph was used as reference monitor for steps, MVPA, and TEE. The Actiheart was used as criterion measure for RHR. The sleep diary (time in bed) was used to compare sleep duration (Polar: sleep time, Oura: total sleep). Normality was tested using the Shapiro-Wilkins test and bootstrapping was used on all variables. Correlations were calculated using repeated measures correlation (RMC) [20-22], with correlation cut-offs suggested by Evans [23]. Mean absolute percentage error (MAPE) was calculated using a cut-off of 10% to indicate low error for studies conducted in free-living [24, 25]. Bland-Altman plots for multiple measurements was created to determine agreement between each instrument and the reference monitor [26].

## Results

### Participant characteristics

Summary statistics for the participants’ age, sex, height, weight, and body mass index, are given in Table 2.

**Table 2.** Participant characteristics for age, sex, height, weight, and body mass index (N=21).

| Variable | Value | Range |
| --- | --- | --- |
| Age, years | 33 (14) | 22-71 |
| Males, % | 57 (12) | N/A |
| Height, cm | 176 (9) | 160-190 |
| Weight, kg | 79 (13) | 57-103 |
| Body mass index, kg/m<sup>2</sup> | 26 (4) | 18-35 |

Values are means (standard deviations) and range, or percentages (numbers). N/A: Not applicable

Each participant had 2-6 valid days of physical activity recording, totalling 57 (Polar Vantage) and 68 (Oura) valid person-days of simultaneous ActiGraph and activity tracker usage. On average each participant simultaneously wore the ActiGraph and Polar Vantage 2.7 days and the ActiGraph and Oura 3.2 days. Participants manually recorded 0-5 days of sleep, totalling 48 (Polar Vantage) and 44 (Oura) person-days of sleep diary and activity tracker recordings. On average each participant kept a sleep diary for 2.3 days. There were 39 person-days of Oura and Actiheart RHR recordings, averaging 1.8 days of recording per participant.

### Correlation and agreement

Table 3 presents valid person-days, correlations, MAPEs, mean difference with limits of agreement for steps, MVPA, TEE, sleep duration, and RHR (Oura), for the Polar Vantage and the Oura, respectively.

**Table 3.**
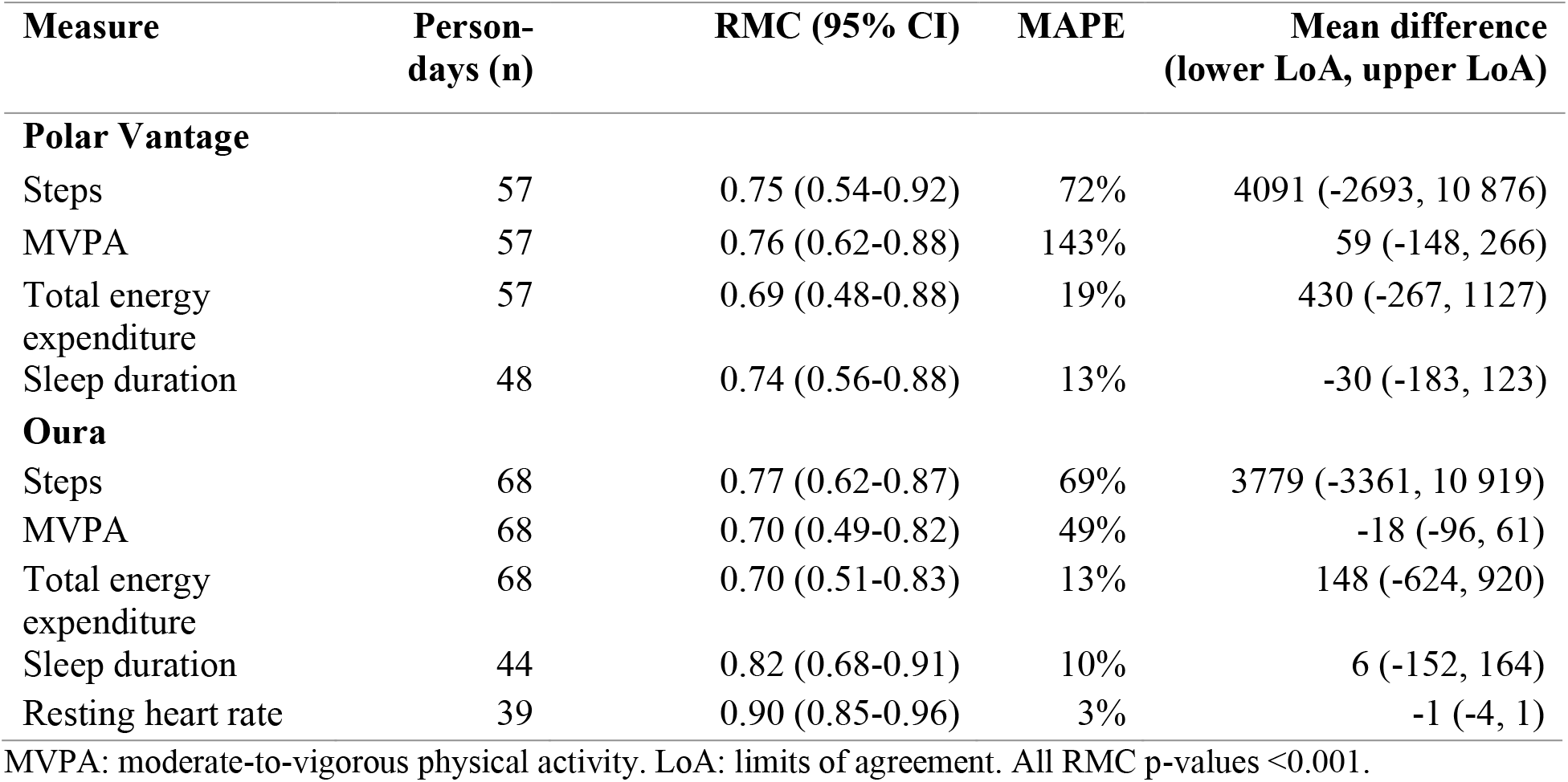
Repeated measures correlation (RMC), mean absolute percentage errors (MAPE), and mean difference for each Polar Vantage and Oura variable.

Numbers are also illustrated in Figure 2 with RMC scatterplots and Bland-Altman plots for both activity trackers. Each participant is represented with a different colour. In the scatterplots, a separate fit-line is shown for each participant.

**Figure 2.**
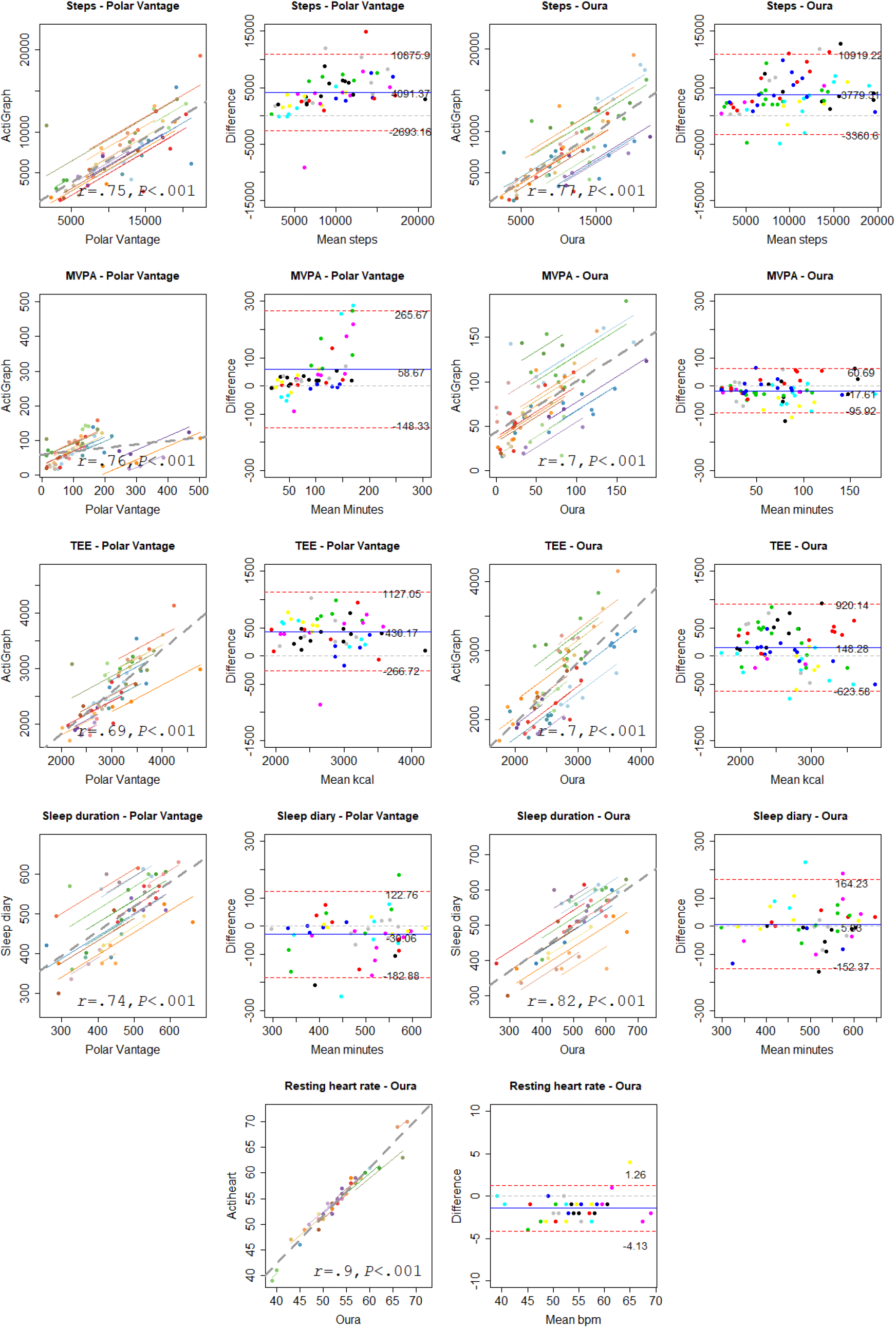
Polar Vantage and Oura scatterplots and Bland-Altman plots for all variables. Each participant’s daily plots and fit lines are represented with a different colour.

*Step* counting was strongly correlated between the reference monitor and both the Polar Vantage and the Oura. Bland-Altman plots showed that both the Polar Vantage and the Oura over-reported steps. The confidence interval range and MAPE were high for both activity trackers.

*MVPA* was strongly correlated between the reference monitor and both activity trackers. The Polar Vantage over-reported MVPA, while the Oura under-reported MVPA. For the Polar Vantage, over-reporting was higher for higher values of MVPA. The limits of agreement range in the Bland-Altman plots was also wider for the Polar Vantage. The confidence interval ranges and MAPEs were high, but the Oura had less mean error compared to the Polar Vantage.

*TEE* was strongly correlated between the reference monitor and both activity trackers. Both activity trackers over-reported TEE, but the Polar Vantage over-reported at a higher rate. The confidence interval ranges were high. Both MAPEs were higher than the 10% cut-off for acceptable error.

*Sleep duration* was strongly correlated between the diary and the Polar Vantage, with a mean under-reporting of 30 minutes by the Polar Vantage. The Oura was very strongly correlated with the diary, but over-reported sleep by on average six minutes. Both activity trackers had a borderline acceptable MAPE, but the confidence interval ranges were high.

*RHR* was very strongly correlated between criterion and the Oura. MAPE was low at 3%, and on average the Oura under-reported RHR with one beat per minute. The confidence interval range was borderline acceptable (11%).

## Discussion

### Principal findings

Step-, MVPA-, and TEE-outputs from the activity trackers were all strongly correlated with the ActiGraph. MAPE was high for these variables, with less error recorded by Oura, compared to Polar Vantage. Sleep duration was strongly to very strongly correlated between the sleep diary and both the Polar Vantage and the Oura. In addition, RHR, recorded by Oura, was very strongly correlated with the Actiheart. MAPEs for the sleep diary were borderline acceptable, while MAPE for RHR was acceptable. The confidence interval range was high for all correlation, with a borderline acceptable range for Oura RHR.

### Strength and limitations

The major strength of this study is the inclusion of multiple days of recordings for each participant instead of one mean measure per participant. A further strength is the gender balance and the wide range in age, height, weight, and body mass index among participants. This is also the first study assessing the accuracy of physical activity estimates from the Oura ring. The major weakness of this study is the use of non-gold standard criteria for comparing most variables. However, the criteria used; the ActiGraph [10-12] and Actiheart [13], have both been previously validated against gold standards, and were found appropriate for this study conducted in free-living.

### Comparison with prior work

While correlations for energy expenditure in the present study are higher than most previous Polar studies (only discontinued devices) [27], it is lower compared to a 2019 lab-setting study on Polar Vantage [8] and a 2018 free-living study on Polar M430 (also discontinued) [28]. Correlations for MVPA on previous Polar validation studies varies, but our findings are similarly high compared to findings from Polar M430 [28] and Polar V800 [29]. Similarly, strong correlations for step counting is in accordance with previous studies on Polar V800 [28, 29] and Polar M600 [30]. For Oura, we could not find any previous studies on steps, MVPA, or TEE. Even though correlations are strong in the present study, most results showed that measurement error was high, and most variables were over-reported compared to the reference monitor. Thus, we cannot recommend replacing existing research-based accelerometers with the activity trackers investigated in the present study. However, as an additional instrument for long term physical activity recording, these consumer-based activity trackers can potentially provide additional value to studies examining changes in physical activity over time. TEE may be especially interesting to measure over time, since it had close to acceptable error, with 13%-19% mean error.

We could not find any previous validation study on sleep duration for the Polar Vantage. However, the high correlation in the present study is in accordance with findings on an earlier Polar model (Polar A370) [4]. Sleep duration was under-reported in both studies, which is expected since sleep onset and sleep offset are generally greater than zero minutes. The Oura over-reported sleep duration, but only by an average six minutes per night. This is in accordance with de Zambotti et al. [9] who conclude that the Oura shows “promising results” for sleep detecting in their study using PSG. Both the Polar Vantage and Oura can provide reasonably close estimates for sleep duration with 10-13% average error, which is close to the acceptable cut-off. This is especially interesting for long term sleep monitoring, as keeping a manual sleep diary is prone to low compliance [7], and objective wrist-worn devices can gives more accurate results [31].

The strong correlation and low average error found for Oura RHR is in accordance with heart rate measurements from wrist-worn activity trackers [5, 32]. A recent validation study on Oura sleeping heart rate and heart rate variability also concluded high agreement between the Oura and an ECG [33]. Compared to the Actiheart, and other similar research instruments, the Oura is a low burden device capable of collecting various heart rate measures over several months. This makes it an interesting instrument for future research, when long-term heart rate measuring is of interest.

Several systematic reviews on consumer-based activity tracker validity have been published the last few years with similar conclusions [27, 32, 34-39]. Fuller et al. [32] published a large systematic review in 2020 assessing the validity of steps, energy expenditure, and heart rate estimates for devices from Apple, Fitbit, Garmin, Mio, Misfit, Polar, Samsung, Withings, and Xiaomi. From 148 included validation studies they concluded that, although there is variation, consumer-based activity trackers can accurately measure steps and heart rate in lab-settings. The most recent systematic review on Garmin activity trackers by Evenson et al. [39] concluded high validity of steps, but low validity for energy expenditure and heart rate. A systematic review and meta-analysis on energy expenditure by O’Driscoll et al. [38] further showed that accuracy is dependent on activity type performed, and that trackers with heart rate monitoring gives lower measurement error.

Although several systematic reviews were released this year [27, 32, 38, 39], all included validation studies were done on activity trackers that are now discontinued. This highlights the need for continuous validation studies on new devices, as suggested by Fuller et al. [32].

## Conclusions

The Oura can be used as an alternative to Actiheart to measure RHR, unless very high accuracy is required. Similarly, Polar Vantage and Oura can potentially be used as a replacement for a manual sleep diary, for sleep duration measurements. Neither the Polar Vantage nor the Oura can replace the ActiGraph for measuring steps, MVPA, and TEE, but may be used as an additional source of physical activity in some settings.

## Data Availability

Data is available from the first author upon reasonable request

## Acknowledgements

AH, FS, and LH conceived the study and designed the study protocol. AH collected data with support from Celina Jakobsen and Jørgen Halvorsen. AH performed data analysis, and AH, LH, SG, GH, and FS contributed to the interpretations of the results. AH drafted the initial manuscript with critical review from LH, SG, FS, and GH. All authors read and approved the final manuscript. The authors would like to thank Celine Jakobsen and Jørgen Halvorsen for their contribution with data collection.

This work was supported by UiT The Arctic University of Norway’s thematic priority grant Personalized medicine for public health (grant Series-1). The publication charges for this article have been funded by a grant from the publication fund of UiT The Arctic University of Norway.

## Ethics Approval

The Norwegian Regional Committees for Medical and Health Research Ethics North reviewed the study (2019/557/REK nord). Participants received written and oral instructions and gave informed consent. The study was conducted in accordance with the 1964 Declaration of Helsinki and its later amendments.

## Abbreviations

ECG: Electrocardiogram
MAPE: Mean absolute percentage error
MVPA: Moderate-to-vigorous physical activity
PSG: Polysomnography
RHR: Resting heart rate
RMC: Repeated measures correlation
TEE: Total energy expenditure

